# Phase-adjusted estimation of the number of Coronavirus Disease 2019 cases in Wuhan, China

**DOI:** 10.1101/2020.02.18.20024281

**Authors:** Huwen Wang, Zezhou Wang, Yinqiao Dong, Ruijie Chang, Chen Xu, Xiaoyue Yu, Shuxian Zhang, Lhakpa Tsamlag, Meili Shang, Jinyan Huang, Ying Wang, Gang Xu, Tian Shen, Xinxin Zhang, Yong Cai

**Author notes:** These authors contributed equally: Huwen Wang, Zezhou Wang, Yinqiao Dong, Ruijie Chang, Chen Xu, Xiaoyue Yu, Shuxian Zhang, Lhakpa Tsamlag. Correspondence: Xinxin Zhang or Yong Cai.

## Abstract

An outbreak of clusters of viral pneumonia due to a novel coronavirus (2019-nCoV / SARS-CoV-2) happened in Wuhan, Hubei Province in China in December 2019. Since the outbreak, several groups reported estimated R_0_ of Coronavirus Disease 2019 (COVID-19) and generated valuable prediction for the early phase of this outbreak. After implementation of strict prevention and control measures in China, new estimation is needed. An infectious disease dynamics SEIR (Susceptible, Exposed, Infectious and Removed) model was applied to estimate the epidemic trend in Wuhan, China under two assumptions of R_t_. In the first assumption, R_t_ was assumed to maintain over 1. The estimated number of infections would continue to increase throughout February without any indication of dropping with R_t_ = 1.9, 2.6 or 3.1. The number of infections would reach 11,044, 70,258 and 227,989, respectively, by 29 February 2020. In the second assumption, R_t_ was assumed to gradually decrease at different phases from high level of transmission (R_t_ = 3.1, 2.6 and 1.9) to below 1 (R_t_ = 0.9 or 0.5) owing to increasingly implemented public heath intervention. Several phases were divided by the dates when various levels of prevention and control measures were taken in effect in Wuhan. The estimated number of infections would reach the peak in late February, which is 58,077–84,520 or 55,869–81,393. Whether or not the peak of the number of infections would occur in February 2020 may be an important index for evaluating the sufficiency of the current measures taken in China. Regardless of the occurrence of the peak, the currently strict measures in Wuhan should be continuously implemented and necessary strict public health measures should be applied in other locations in China with high number of COVID-19 cases, in order to reduce R_t_ to an ideal level and control the infection.

## Introduction

2019 novel coronavirus (2019-nCoV / SARS-CoV-2) has given rise to an outbreak of viral pneumonia in Wuhan, China since December 2019.^1,2^ World Health Organization (WHO) now has named the disease COVID-19, short for “coronavirus disease 2019”.^3^ Most cases from the initial cluster had an epidemiological link to the Huanan Seafood Wholesale Market.^4^ Patients have clinical manifestations including fever, cough, shortness of breath, muscle ache, confusion, headache, sore throat, rhinorrhoea, chest pain, diarrhoea, and nausea and vomiting.^5,6^ As of 17 February 2020, a cumulative total of 72,436 confirmed cases (including 11,741 currently severe cases), 6,242 currently suspect cases, a cumulative total of 1,868 deaths and 12,552 cases discharged from hospital were reported by National Health Commission of the People’s Republic of China (NHC) in mainland China.^7^ The significant increases in the number of confirmed cases in China and abroad led to the announcement made by WHO on 30 January that the event has already constituted a Public Health Emergency of International Concern.^8^

The reproduction number, R, measures the transmissibility of a virus, representing the average number of new infections generated by each infected person, the initial constant of which is called the basic reproduction number, R_0_,^9^ and the actual average number of secondary cases per infected case at time t is called effective reproduction number, R_t_.^10-12^ R_t_ shows time-dependent variation with the implementation of control measures. R >1 indicates that the outbreak is self-sustaining unless effective control measures are implemented, while R <1 indicates that the number of new cases decreases over time and, eventually, the outbreak will stop.^9^ Over the past month, several groups reported estimated R_0_ of COVID-19 and generated valuable prediction for the early phase of this outbreak. In particular, Imai et al.^9^ provided the first estimation, using R_0_ of 2.6 and based on the number of cases in China and those detected in other countries. Other authors estimated R_0_ to be 3.8,^13^ 6.47,^14^ 2.2,^15^ and 2.68.^16^ These predictions were very alerting and suggestions have been made for very strict public health measures to contain the epidemics.

In response to the outbreak of COVID-19, a series of prompt public health measures have been taken. On 1 January, the Huanan Seafood Wholesale Market was closed in the hope of eliminating zoonotic source of the virus.^5^ On 11 January, upon isolation of the viral strain for COVID-19 and establishment of its whole genome sequences,^17^ RT-PCR reagents were developed and provided to Wuhan, which ensured the fast ascertainment of infection.^15^ On 21 January, Emergency Response System was activated to better provide ongoing support to the COVID-19 response.^18^ Ever since the outbreak, the work of intensive surveillance, epidemiological investigations and isolation of suspect cases gradually improved. Those having had close contacts with infections were asked to receive medical observation and quarantine for 14 days.^19^ Travel from and to Wuhan City as well as other medium-sized cities in Hubei Province has been restricted since 23 January 2020.^20^

The 2019-nCoV / SARS-CoV-2 has at least 79.5% similarity in genetic sequence to SARS-CoV.^5,17^ Riley estimated that 2.7 secondary infections were generated per case on average (R_0_= 2.7) at the start of the SARS epidemic without controlling.^21^ After isolating the patients and controlling the infection by the authority, the value of R_t_ for SARS decreased to 0.25.^22^ As Li et al. mentioned, it is possible that subsequent control measures in Wuhan, and elsewhere in mainland China, have reduced transmissibility.^15^ A new estimation of the epidemic dynamics taking the unprecedentedly strict prevention and control measures in China into consideration is required to better guide the future prevention decisions.

In this article, we intended to make phase-adjusted estimation of the epidemic trend for the 2019-nCoV / SARS-CoV-2 infection transmission in Wuhan, China under two assumptions of R_t_ (maintaining high >1 or gradually decreasing to <1). We hope to depict two types of epidemic dynamics to provide potential evaluation standard for the effects of current prevention and control measures, and to provide theoretical basis for future prevention decisions of the current epidemic in China.

## Results

### Estimation of the epidemic trend assuming that the prevention and control measures are insufficient in Wuhan, China

Assuming the epidemic continues to develop with R_0_ = 1.9, 2.6 and 3.1^9^ from 1 December 2019, the number of infections will continue to rise (Fig. 1). By the end of February 2020, COVID-19 cases would be 11,044, 70,258 and 227,989 in Wuhan, China with R_0_ = 1.9, 2.6 and 3.1, respectively. Detailed calculation process is included in the Materials and Methods part.

**Fig. 1.**
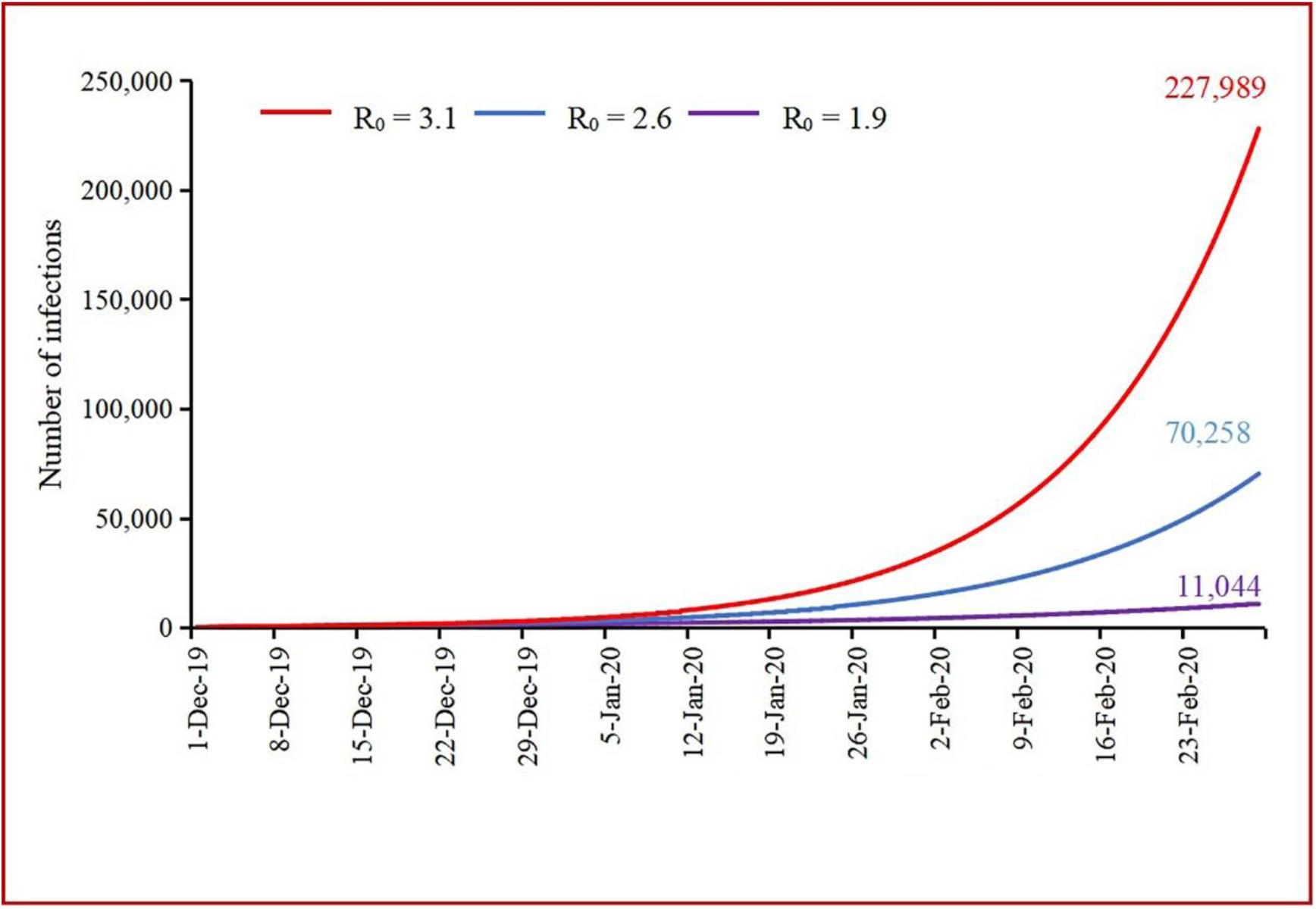
Estimation of the number of COVID-19 cases in Wuhan, China (December 2019– February 2020, R_0_ = 1.9, 2.6 and 3.1). 11,044, 70,258 and 227,989 represent the estimated number of COVID-19 cases by the end of February 2020 in Wuhan, China with R_0_ = 1.9, 2.6 and 3.1, respectively.

### Estimation of the epidemic trend assuming that the prevention and control measures are sufficient in Wuhan, China

The first phase (1 December 2019–23 January 2020): It was the early phase of the epidemic when a few prevention and control measures were implemented. The number of infections in Wuhan, China reached 17,656–25,875 by the end of this phase with R_0_ as 3.1.

The second phase (24 January 2020–2 February 2020): From 23 January 2020 on, public transportations to and from Wuhan, as well as public transportation within Wuhan were stopped. While gathering events inside Wuhan was banned, quarantine and isolation were gradually established in Wuhan. The number of infections was 32,061–46,905 by the end of this phase as R_t_ decreased to 2.6.

The third phase (3 February 2020–15 February 2020): New infectious disease hospitals and mobile cabin hospitals came into service and many medical and public health teams from other provinces and cities in China arrived in Wuhan. The quarantine and isolation at community level were further enhanced. The number of infections would reach 53,070–77,390 if R_t_ could be reduced sequentially to 1.9.

The fourth phase (from 15 February 2020 on): All of the most restrict public health measures may need a longest incubation period to take effect. If R_t_ could be gradually reduced to 0.9 or 0.5 in the fourth phase, the epidemic peaks and inflection points might occur in Wuhan, China on 23 February or 19 February. The number of infections would be 58,077–84,520 or 55,869–81,393 with R_t_ = 0.9 or 0.5, respectively (Figs. 2-3).

**Fig. 2.**
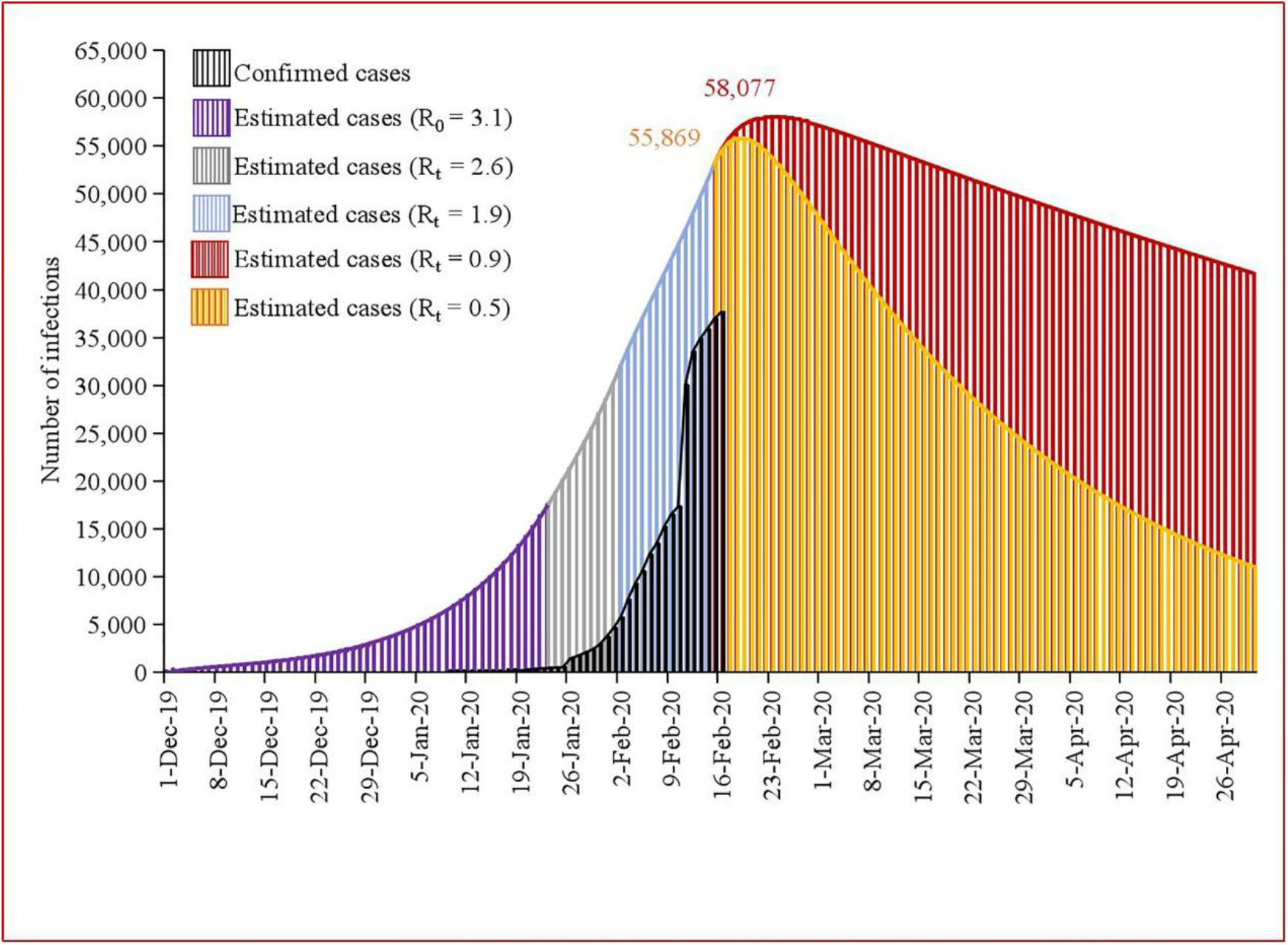
Phase-adjusted estimation of the number of COVID-19 cases in Wuhan, China (1 December 2019–30 April 2020, E = 20I). 55,869 represents the estimated peak number of COVID-19 cases on 19 February 2020 in Wuhan, China with R_0_ = 0.5; 58,077 represents the estimated peak number of COVID-19 cases on 23 February 2020 in Wuhan, China with R_0_ = 0.9; E: number of exposed cases; I: number of infectious cases; E was assumed to be 20 times of I at baseline.

**Fig. 3.**
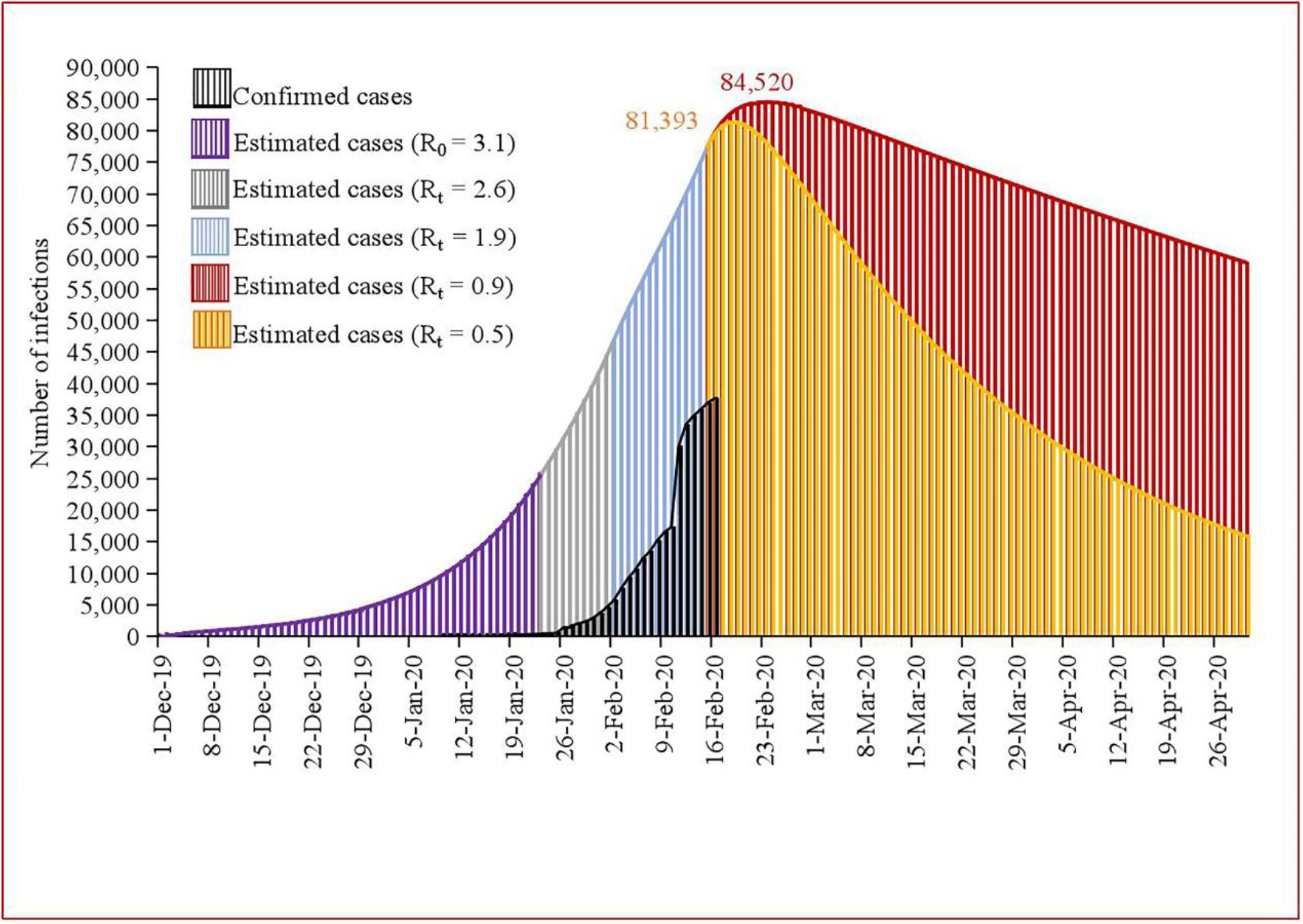
Phase-adjusted estimation of the number of COVID-19 cases in Wuhan, China (1 December 2019–30 April 2020, E = 30I). 81,393 represents the estimated peak number of COVID-19 cases on 19 February 2020 in Wuhan, China with R_0_ = 0.5; 84,520 represents the estimated peak number of COVID-19 cases on 23 February 2020 in Wuhan, China with R_0_ = 0.9; E: number of exposed cases; I: number of infectious cases; E was assumed to be 30 times of I at baseline.

Our model predicted 2,323–3,381 deaths in Wuhan, China when we assumed R_t_ as 0.9 and the percent of deaths as 4%; 2,235–3,256 deaths when we assumed R_t_ as 0.5 at the fourth phase. An average of 2,279–3,318 deaths were also estimated (Table 1).

**Table 1.** Estimating the number of deaths of COVID-19 cases in Wuhan, China (R_t_ = 0.9 or 0.5)

| | $R_t = 0.9$ | $R_t = 0.5$ | Average |
| --- | --- | --- | --- |
| Total cases | 58,077–84,520 | 55,869–81,393 | 56,973–82,957 |
| Deaths (4%) | 2,323–3,381 | 2,235–3,256 | 2,279–3,318 |
| Deaths (10%) | 5,808–8,452 | 5,587–8,139 | 5,697–8,296 |

The estimated percent of deaths is about 4%–10%.<sup>6,24</sup>

When we assumed R_t_ as 0.9 and the percent of deaths 10% based on calculation of case fatality rate (CFR) at early stage of the epidemic,^6^ our model predicted 5,808–8,452 deaths in Wuhan, China; 5,587–8,139 deaths when we assumed R_t_ as 0.5 at the fourth phase. An average of 5,697–8,296 deaths were also estimated.

## Discussion

Estimations of the transmission risk and the epidemic trend of COVID-19 are of great importance because these can arouse the vigilance of the policy makers, health professionals and the whole society so that enough resources would be mobilized in a speedy and efficient way for both control and treatment. We estimated the number of infections using SEIR model under two assumptions of R_t_ (R_t_ maintaining to be >1 or R_t_ gradually decreasing to <1) in the purpose of depicting various possible epidemic trends of COVID-19 in Wuhan, China. Two estimations provide an approach for evaluating the sufficiency of the current measures taken in China, depending on whether or not the peak of the number of infections would occur in February 2020. Assuming the current control measures were ineffective and insufficient, the estimated number of infections would continue to increase throughout February without a peak. On the other hand, assuming the current control measures were effective and sufficient, the estimated number of infections would reach the peak in late February 2020.

According to Read’s research,^13^ R_0_ for COVID-19 outbreak is much higher compared with other emergent coronaviruses. It might be very difficult to contain or control the spreading of this virus. If the prevention and control measures were not sufficient or some new factors occurred (e.g. a large proportion of cases with mild or none symptoms existed in the community; there were more zoonotic sources), the epidemic might continue to develop at a high speed. Therefore, we depicted first the epidemic dynamics of the relatively unsatisfying circumstance based on the R_0_ estimated before the unprecedented efforts of China in the containment of the epidemics occurred and the newest documented parameters. The curve continued to go up throughout February without any indication of dropping, indicating the need for further enhancement of public health measures for containment of the current outbreak.

However, as mentioned by WHO in the statement on 30 January, “it is still possible to interrupt virus spread, provided that countries put in place strong measures to detect disease early, isolate and treat cases, trace contacts, and promote social-distancing measures commensurate with the risk.”^8^ Responding to the outbreak, China has taken a series of unprecedentedly strict measures regardless of economic losses including daily contact with WHO and comprehensive multi-sectoral approaches to fight against the virus and prevent further spread, showing the sense of responsibility of China to its citizens and the whole world. Epidemic information has been released in an open, transparent, responsible and timely manner home and abroad. Cooperation has been established with other countries and international organizations. These measures have won full recognition of the international community, including WHO. Specifically in Wuhan, in the early phase from beginning of December 2019 to 23 January 2020, there was no limitation of population flow and gathering. When the human-to-human transmission was confirmed, an important decision was made to isolate Wuhan from other parts of the country. As a result, since 24 January 2020, all public transports from and to Wuhan, as well as public transports and people’s gathering events within Wuhan, were stopped. Since 2 February 2020, strict public health measures were taken to prevent population flow among distinct communities whereas since 9 February 2020, public health interventions including quarantine of each building in the urban area and each village in the rural area were implemented in order to block the transmission chain among the household. Therefore, strong efforts of authorities and people in Wuhan with the support of the central government and people from all over China, as well as the WHO and the international society, may have gradually braked COVID-19 outbreak.

R_t_ is therefore assumed to decrease gradually from 3.1 to 0.5 in Wuhan, China in the current study. The trend of the estimated cases is in accordance with the trend of currently confirmed cases. The relatively big difference in number may be due to the possible existence of a large number of mild and asymptomatic cases and the imperfection of current diagnostic measures. According to NHC, before 12 February 2020, the confirmed cases were diagnosed according to contact history, clinical manifestations, chest X-ray or computer assisted tomography (CT) and RT-PCR for COVID-19. Since 12 February 2020, the diagnosis has been mainly based on contact history, clinical manifestation and imaging evidence of pulmonary lesion suggestive of pneumonia, while viral detection with RT-PCR is still being performed in a part of patients.^23^ After the diagnosis method was changed, a large number of cases that were previously missed and piled up for testing were reported in Wuhan, which greatly increased the number of existing cases and made it approaching our estimated number. A peak of the estimated number of infections would occur in late February under this assumption. If the peak does occur in February, the very strong measures China has taken may have already received success in controlling the COVID-19 infection in Wuhan.

The number of deaths in the current study was estimated based on previously reported CFRs. Chen et al. calculated it to be 11% based on 99 cases at the very early stage of the outbreak.^6^ This mortality rate might not be representative of the whole patients population due to the relatively small sample size and scarce knowledge about the virus at early stage. More recently, Yang et al. estimated the overall adjusted CFR among confirmed patients to be 3.06% with a sample size of 8,866.^24^ The number of deaths estimated accordingly might be more close to the reality. Our estimation of number of deaths only provides a possible range based on currently reported CFR. The actual number of deaths might be lower with more mild and asymptomatic cases being detected and the improvement of clinical care and treatment as the epidemic progresses.

Hubei Province, of which Wuhan is the capital city, accounts for more than 80% of newly-confirmed cases all over the country according to NHC daily report. The current epidemic trend in Hubei Province is similar to that in Wuhan City. Considering the high number of confirmed cases in the province, the currently strict measures should be continuously implemented both in Wuhan and other cities in Hubei Province no matter whether the peak of number of infections would occur or not, in order to reduce R_t_ to an ideal level and to control the epidemic. Owing to the timely transportation restriction in Hubei Province and other measures, the number of newly-confirmed cases remains relatively low and has decreased for 13 days in a row in other provinces, autonomous regions and municipalities in mainland China outside Hubei Province. However, independent self-sustaining human-to-human spread is estimated to already present in multiple major Chinese cities including Beijing, Shanghai, Guangzhou, and Shenzhen.^16^ In addition, pressure on transmission control caused by the population migration after Spring Festival holidays may occur soon, especially in some densely-populated cities.^25^. Necessary strict measures should still be maintained even when the current measures turn out to be effective.

Our study has some limitations. Firstly, the SEIR model was set up based on a number of necessary assumptions. For example, we assumed that no super-spreaders exist in the model, but there is currently no supportive evidence. Secondly, the accuracy of the estimation model depends largely on the accuracy of the parameters it used such as incubation period. With more precise parameters obtained as the epidemic progresses, our estimation model will also be more precise. Our estimates of the reproductive number from 3.1 to 0.5 are based on previous studies and experience from SARS control. However, this measure may change substantially over the course of this epidemic and as additional data arrives. Besides, using a fixed R_t_ value in each phase may incur potential bias because R_t_ is essentially a dynamic parameter. Thirdly, these estimated data may not be sustained if unforeseeable factors occurred. For example, if some infections were caused by multiple exposures to animals, these estimates will be exposed to a big uncertainty. Fourthly, the epidemic trend shows great difference between Wuhan and Hubei Province and regions in mainland China outside Hubei Province according to the NHC reported data. It is thus inappropriate to generalize the estimations in Wuhan to regions in mainland China outside Hubei Province. The dynamics model for the other locations in mainland China remains to be developed and specific parameters need to be redefined. Lastly, we do not provide model fit information in the current study. SEIR model is a prediction model forecasting the number of infections in the future. The data corresponding to actual situation in the future cannot be determined and this makes model fitting almost impossible during the outbreak. We would carry out model fitting according to the real data in pace with more information and knowledge about the characteristics of COVID-19 and the epidemics in the future.

Despite the limitations mentioned above, the current study is the first to provide estimation for epidemic trend after strict prevention and control measures were implemented in China. Whether current prevention and control measures are sufficient or not may be evaluated through the occurrence of the infection number peak in February. Rigorous measures should still be maintained even when the current measures turn out to be effective by the end of February to prevent further spread of the virus.

## MATERIALS AND METHODS

### Model

We employed an infectious disease dynamics model (Susceptible, Exposed, Infectious and Removed model; SEIR model) for the purpose of modeling and predicting the number of COVID-19 cases in Wuhan, China. The model is a classic epidemic method to analyze the infectious disease which has a definite latent period, and has proved to be predictive for a variety of acute infectious diseases in the past such as Ebola and SARS.^22,26-31^ Application of the mathematical model is of great guiding significance to assess the impact of isolation of symptomatic cases as well as observation of asymptomatic contact cases and to promote evidence-based decisions and policy.

We assumed no new transmissions from animals, no differences in individual immunity, the time-scale of the epidemic is much faster than characteristic times for demographic processes (natural birth and death), and no differences in natural births and deaths. In this model, individuals are classified into four types: susceptible (S; at risk of contracting the disease), exposed (E; infected but not yet infectious), infectious (I; capable of transmitting the disease), and removed (R; those who recover or die from the disease). The total population size (N) is given by N = S+E+I+R. It is assumed that susceptible individuals who have been infected first enter a latent (exposed) stage, during which they may have a low level of infectivity. The differential equations of the SEIR model are given as:^32,33^

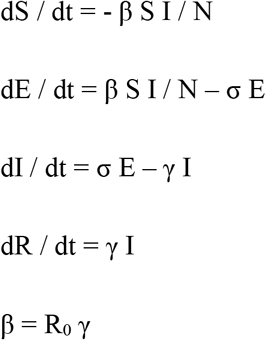

where β is the transmission rate; σ is the infection rate calculated by the inverse of the mean latent period; γ is the recovery rate calculated by the inverse of infectious period.

R software (Version 3.6.2) was applied for all the calculations and estimates in the current study.

### Data collection and parameter values

#### Estimation of the epidemic trend assuming the prevention and control measures are insufficient

We first estimated the epidemic trend in Wuhan, China assuming the current prevention and control measures are insufficient. In this process, S was assumed to be the population of Wuhan City (11 million).^15,34^ The initial assumed number of cases caused by zoonotic exposure was 40 (I) according to Imai et al.’s estimation.^9^ We proposed E at 20 times of I in accordance with Read et al.^13^ R was set as 0. σ was set as 1/5.2 according to the latest article by Li et at.^15^, which calculated the mean incubation period of COVID-19 to be 5.2 days. Chen et al.^6^ calculated the average hospitalization period of 31 discharged patients to be 12.39 ± 4.77 days. Yang et al. calculated the median time from disease onset to diagnosis among confirmed patients to be 5.^24^ γ was accordingly set as 1/18 (ceiling of 12.39 + 5 is 18). R_0_ was chosen based on Imai et al.’s^9^ estimation 2.6 (1.9–3.1) assuming 4,000 (1,000–9,700) infections as of 18 January.

#### Estimation of the epidemic trend assuming the prevention and control measures are sufficient

This section discussed the scenario where the current prevention and control measures are sufficient. The set of S, E, I, R, σ and γ is the same as the first section except that we also explored the model with E at 30 times of I to provide a possible range of number of infections. Absence of fever in COVID-19 cases is more frequent than in SARS-CoV and MERS-CoV infection.^35^ Such patients may be missed since the current surveillance case definition focused mainly on fever detection. Accordingly, the possibility of E at 30 times of I can’t be excluded.

R_0_ in this section was chosen by phases. The first phase ranges from 1 December 2019 to 23 January 2020 and can be regarded as the early phase of the epidemic when a few prevention and control measures were implemented. R_0_ was set as 3.1 consistent with Imai et al.’s estimation of high transmission level.^9^ On 23 January 2020, airplanes, trains and other public transportation within the city were restricted and other prevention and control measures such as quarantine and isolation were gradually established in Wuhan.^20^ So the second phase began on 24 January and R_t_ was set as 2.6 consistent with Imai et al.’s estimation of moderate transmission level.^9^ 2 February was the last day of the extended Spring Festival holiday and Chinese authorities mobilized more medical resources to support Wuhan ever since.^36^ The newly constructed hospital “Huoshenshan” come into service on this day^37^ and “Leishenshan”, mobile cabin hospitals several days later.^38^ And also, more and more medical teams arrived in Wuhan. So the third phase began on 3 February and R_t_ was set as 1.9 consistent with Imai et al.’s estimation of low transmission level.^9^ All of these measures may need one longest incubation period to take effect. So the last phase began on 16 February and R_t_ was set as 0.9 and 0.5, respectively, assuming the prevention and control measures are sufficient and effective to depict two different levels of effect of the measures in reducing transmission probability.

## Data Availability

The data used in the paper all come from official reports.

## Acknowledgments

This work is funded by Medicine and Engineering Interdisciplinary Research Fund of Shanghai Jiao Tong University (YG2020YQ06), the National Key Research and Development Project (2018YFC1705100, 2018YFC1705103) and the National Natural Science Foundation of China (71673187), which had no role in study design, data collection, data analysis, data interpretation, or writing of the report. We acknowledge all health-care workers involved in the diagnosis and treatment of patients all around China. We thank National Health Commission of the People’s Republic of China for coordinating data collection for patients with COVID-19 infection.

## Contributions

YC, XZ, HW, ZW, YD had the idea for and designed the study and had full access to all data in the study and take responsibility for the integrity of the data and the accuracy of the data analysis. YC, HW, ZW, YD, RC, SZ, CX, LT and XY contributed to writing of the manuscript. HW, YC, JH, GX, TS, YW, XZ contributed to critical revision of the report. YC, XZ, HW, ZW, YD, MS, RC, SZ, CX, LT, GX, TS, YW and XY contributed to the statistical analysis. All authors contributed to data acquisition, data analysis, or data interpretation, and reviewed and approved the final version.

The current manuscript was accepted by Cell Discovery on 18 February 2020.

